# THREE STEPS NOVEL HARD MARGIN ENSEMBLE MACHINE LEARNING METHOD CLASSIFIES UNCERTAIN *MEFV* GENE VARIANTS

**DOI:** 10.1101/2023.04.08.23288306

**Authors:** Mustafa Tarık Alay, İbrahim Demir, Murat Kirişçi

## Abstract

**Introduction:** The International Study Group for Systemic Autoinflammatory Diseases (INSAID) consensus criteria revealed that the clinical outcomes of more than half of the MEFV gene variants are uncertain. We aimed to detect more accurate classifications of MEFV variants while simultaneously reducing MEFV variant uncertainty.

**Material-Methods:** We extracted variants of the MEFV gene from the infevers database. We then determined the optimal number of in silico instruments for our model. On the training dataset, we implemented seven machine learning algorithms on MEFV gene variants with known clinical effects. We evaluated the effectiveness of our model in three steps: First, we performed machine-learning algorithms on the training dataset and implemented those with a prediction accuracy of greater than 90 percent. Second, we compared our gene-level and protein-level prediction results. Finally, we compared our prediction results to clinical outcomes.

**Results:** Our analysis included 266 of 381 MEFV gene variants and four computational tools (Revel, SIFT, MetaLR, and FATHMM). In our training dataset, the accuracy of three machine learning algorithms (RF: 100%, CRAT: 100%, and KNN: 91%) exceeded the threshold value. Thus, the dataset contained 134 likely pathogenic (LP) variants and 132 likely benign (LB) variants. We found that B30.2 domain variants were 2.5 times more likely to be LP than LB (χ2:12.693, p < 0.001, OR: 2.595 [1.532-4.132].

**Discussion:** Considering that the clinical effects of 60% of MEFV gene variants have not yet been determined, a combined evaluation of our methods and patients’ clinical manifestations significantly simplifies the interpretation of unknown variants.

## 1. INTRODUCTION

Familial Mediterranean Fever (FMF) is an inherited autoinflammatory disease characterized by recurrent fever, abdominal pain, and polyserositis^1,2^. High rates of FMF are reported among Turks, Armenians, Middle Easterners, and Jews, with 1 case per 500-700,000 individuals. ^3^. The disease is caused by a mutation in the pyrin protein, which is encoded by the *MEFV* gene. The pyrin protein (marenostrin or TRIM20), which is predominantly expressed in myeloid cells, synovial fibroblasts, and dendritic cells, is composed of 781 amino acids and demonstrates its effects on the innate immune system through the regulation of GTPases and other proteins^4^. Following the activating agent, the mutated pyrin protein remained less phosphorylated and, consequently, more active pyrin propagates inflammation by constructing the pyrin inflammasome. Secretion of interleukine and other inflammatory cytokines, neutrophilia chemotaxis, and induction of an attack by FMF are the subsequent steps^5,6^.

The extensive application of next-generation sequencing technology improves the likelihood of FMF diagnosis, identifies the population’s carrier rates, and predicts the disease’s recurrence risks. However, as an unwanted result of the extensive use of NGS assays, numerous new variations with uncertain clinical significance in *MEFV* gene have been identified^7^. The International Study Group on Systemic Autoinflammatory Disorders (INSAID) consensus criteria found that the clinical outcomes of more than half of the *MEFV* gene variations are uncertain^8^, which is called a variation of uncertain significance (VOUS) for American College of Medical Genetics (ACMG). For example, the clinical impact of 192 out of 358 *MEFV* variations (55.3%) in the infevers database (Infevers: an online database for autoinflammatory mutations) is unknown. Copyright. Accessible at https://infevers.umai-montpellier.fr/web (last access: 05/04/2022)

Interpreting and understanding the clinical effects of VOUS variants is challenging for physicians and patients. Studies implemented in reducing variant uncertainties are most likely based on one ML algorithm, and amino acid prediction score^9–11^. Yet, ensemble algorithms produce more accurate results^12^, although no ensemble method has been applied to classify MEFV gene variants. Well-designed functional and hereditary studies could determine the clinical impact of the VOUS variant, but they are costly and time-consuming; rapid, inexpensive, and risk-free novel methods are required to anticipate the effects of *MEFV* variants^13,14^. Contradictory opinions exist on which and how many protein prediction methods and meta-predictors are employed for clinical variant evaluation^13^. ACMG and Clingen recommend investigating a few specific genes or diseases^15–19^; nevertheless, this is a far cry from predicting the clinical impact of the majority of genes. Novel methods need to be implemented to reduce the uncertainties in clinical evaluation.

In this study, we built a novel tool for more accurate classification of MEFV gene variations by using the optimal number of amino acid prediction scores and machine-learning algorithms. Our goal was to determine a more precise categorization of MEFV variants while also reducing the uncertainties. Our results will assist clinicians in determining the clinical relevance of variations with uncertain clinical impact and in generating gene-specific interpretation guidelines.

## 2. MATERIAL-METHODS

### 2.1. Database accession and Study Design

Initially, we extracted the REVEL, MetaLR, SIFT, and FATHMM scores for 389 MEFV missense variants (deposited at infevers (https://infevers.umai-montpellier.fr/web, last access date:05/04/2022)). Variants with point mutations (missense, silent, and nonsense) in the coding region were included in the study, but other variants (termination gain, termination loss, insertion, frameshift/inframe deletions, indels) were excluded. A study included a total of 266 variants: 3 BENIGN(B), 48 LIKELY BENIGN, 44 LIKELY PATHOGENIC, 5 PATHOGENIC, 110 VARIATION OF UNKNOWN SIGNIFICANCE (VOUS), 26 NOT CATEGORIZED (NC), and 32 UNSOLVED (US).

Therefore, we did not anticipate a batch size effect for any of the infevers variants.

### 2.2. In silico tool selection

During the process of selecting in-silico tools, we evaluated a number of conditions. First, we chose in silico tools because they were up-to-date, validated, and recommended by international guidelines. Second, we meticulously determined that missing values for relevant variant scores in the entire dataset should not be included. Thirdly, we obtained scores from dbNFSP 4.0 and discovered that all variants were utilized by variant data discovery tools such as varsome/franklin. Fourth, we considered the absence of multicollinearity between the chosen scores.

#### 2.2.1. Aminoacid alterations pathogenicity prediction scores

##### SIFT

Sorting intolerant from tolerant(SIFT) aims to differentiate neutral and harmful variants from each other. SIFT sortes all variants from damaged, the effects of amino acid alterations harmful, to tolerable, the effects of amino acid alterations tolerable, and displays better performance in diverse sequences^20^.

##### FATHMM

FATHMM predicts the functional effects of non-synonymous variants by Hidden Markov Models ^21^. The unique disease-system approach of FATHMM provides convenience in reducing false positives rates by discriminating between disease-associated and neutral variants ^22^.

#### 2.2.2. Metapredictors

##### Revel

Most of the tools are trained on neutral variants. However, disease-causing variants most likely stem from rare variants. Revel is an ensemble score that predicts better performance in rare variants which are allele frequency (AF) < 3%^23^.

##### MetaLR

MetaLR is an ensemble predictor which displays better performance in AF>3% and AF <5% than REVEL scores ^23^.

### 2.3. Machine Learning Methods

#### 2.3.1. Selection of Machine learning Methods

In our study, we applied seven machine learning techniques to four scores (SIFT, FATHMM, Revel, and MetaLR): K-nearest neighbor (KNN), Classification and Regression Tree (CART), Random Forest (RF), Logistic regression (LR), Linear Support Vector Machine (LSVM), Polynomial Support Vector Machine (PSVM), and Kernel Support Vector Machine (K-LSVM) (KSVM). Our study only included four features, so we did not employ feature selection techniques such as recursive feature elimination. In addition, prediction scores in our study, with the exception of SIFT, were distributed between 0 and 1; we did not normalize or standardize our algorithms. In addition, RF or CART algorithms naturally do not require feature scaling.

#### 2.3.2. Validation and Evaluation of Machine Learning Algorithms

Our dataset consisted of a small number of samples (n=266), which may have resulted in overfitting in our training dataset. For limited data size, there were two options: increasing the sample size or augmenting the data. Our sample size was small and we were unable to increase it due to the limited number of MEFV variants discovered in the human genome. To overcome this limitation, we selected a data augmentation strategy. Consequently, we employed multiple algorithms in our model; as a result, each score was evaluated four times.

We trained RF, CRAT, KNN, LR, LSVM, KSVM, PSVM on four scores (REVEL, MetaLR, SIFT, FATHMM) and further validated using stratified 5-fold cross-validation. Thus, we were not only reducing overfitting, but also found optimal values of hyperparameters to improve the model efficacy. We used accuracy, precision-recall, and F1 score for evaluation metrics. However, as our training dataset comprised a balanced dataset, we determined our threshold value according to accuracy score. Therefore, we accepted threshold value as 90%. ML algorithms higher than this threshold were used for prediction value. Our prediction scores evaluation was based on emsemle models which was named voting prediction. While implementing voting prediction we determined hard margin software.

### 2.4. Functional and Clinical Evaluation

We evaluated each variant in two categories for functional-level ascertainment: gene-level and protein-level. While we established gene-level evaluation based on exonic position, implemented protein-level evaluation by comparison of pyrin protein domain distributions. We evaluated MEFV domains initiation and termination location according to protein databank (https://www.rcsb.org/), ensemble, interpro (https://www.ebi.ac.uk/interpro/), conserved domain databases (https://www.ncbi.nlm.nih.gov/Structure/cdd/cdd.shtml), prosite (https://www.expasy.org/resources/prosite), and existing literature^24^. *MEFV* (NM_000243.3) transcripts were based on variants distribution on pyrin protein. Clinical level evaluation is restricted by internationally accepted consensus 14 variants with existing literature (Pubmed, Scopus, Web of Science, DOAJ, Google Scholar) and clinical databases(Clinvar, intervar, franklin, varsome, ensembl)^25^.

### 2.5. Statistical Analysis

We conducted a statistical analysis with Python 3.7.1 and SPSS 25.0 for Windows (IBM, Chicago, IL). We checked for normality for both discrete and continuous numerical variables. If assumptions of normality were provided, we considered means and standard deviations. Otherwise, we used medians and interquartile ranges. We expressed all categorical variables (nominal or ordinal) via numbers and percentages and sorted ordinal variables by their hierarchical placement.

We tested assumptions of normality in both graphical and analytical methods (Q-Q plot, detrended plot, boxplot, histogram, and steam-leaf methods for graphical methods, Kolmogorov-Simirnov test for analytical method). Skewness and kurtosis values, of -1 to +1 range, and skewness and kurtosis indexes (skewness and kurtosis divided by standard error), if -2 to +2 range, normality assumptions were provided, otherwise not.

We conducted an Analysis of Variance Test (ANOVA) for normally distributed values. After ANOVA, we performed the Tukey test as a post hoc test. We carried out Kruskall Wallis H-test for non-normally distributed values. After the Kruskall Wallis-H test, we performed the Dunn-Bonferroni test as a post hoc test. The confidence interval determined 95 % for all statistical analyses, with (α) and (β) values accepted as 0.05 and 0.20, respectively. P values threshold defined as 0.05, smaller than this value, were accepted as statistically significant.

## 3. RESULTS

### 3.1. REVEL, SIFT, MetaLR, and FATHMM Prediction Score Distribution for *MEFV* Variants

First, we examined the normality distribution of all four scores and determined that three cases were right-skewed and one was left-skewed. On our dataset, we therefore implemented square root and logarithmic transformations. The Kruskall-Wallis-H test revealed that the dataset remains to have a non-normal distribution for all four scores. Next, we verified whether REVEL, SIFT, MetaLR, and FATHMM scores for the classes “likely benign” vs. “benign”, and “likely pathogenic” vs. “pathogenic” were significantly different. In no cases, the medians of the two benign and two pathogenic classification groups demonstrated statistically significant differences (Kruskal-Wallis test not significant for non-parametric ANOVA of “likely pathogenic” vs. “likely pathogenic/pathogenic” vs. “pathogenic” and “benign” vs. “likely benign”). Therefore, in the remaining statistical analyses, the *MEFV* variants classified at infevers as “Benign” and “Likely Benign” were combined into the category “Likely Benign” (LB). Similarly, the “Likely Pathogenic” and “Pathogenic” classes, were merged into the Likely Pathogenic” (LP) category. These two “LP” and “LB” variants were the features in the training dataset. As the remaining datasets’ clinical outcomes are indeterminate all three categories (Variants of uncertaın significance, Not categorized, Unsolved) merged into Variants of Unkown Significance (VOUS) and used for prediction. The scores distribution and box-swarm plots were shown in Figure 1 and Figure 2, respectively. LP and LB variants were used for the training dataset.

**Figure 1.**
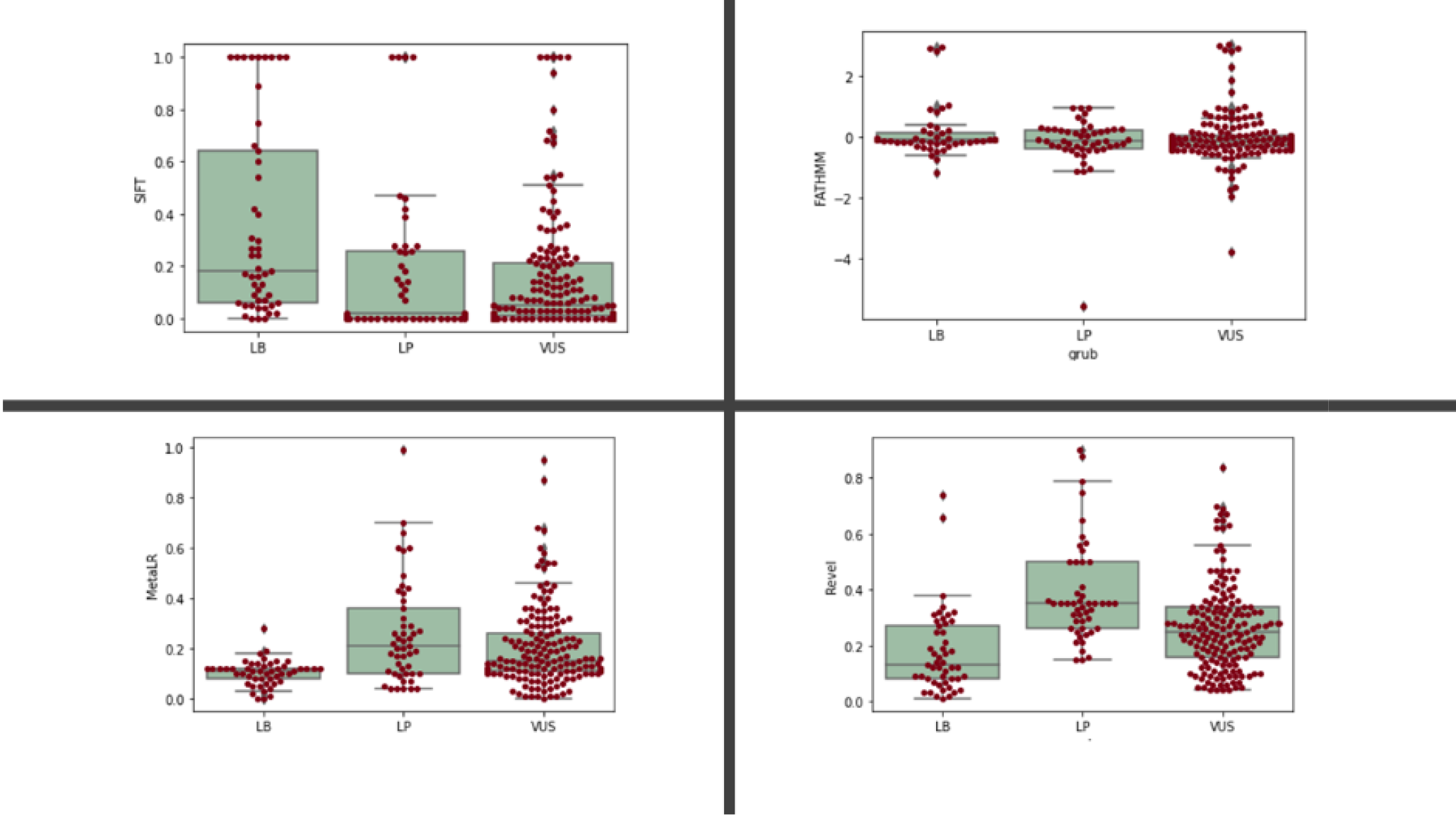
Boxplot and swarmplot for each category. Mann Whitney-U test were performed for each category. Dunn-Bonferronni test implemented for *post hoc* analysis.

**Figure 2.**
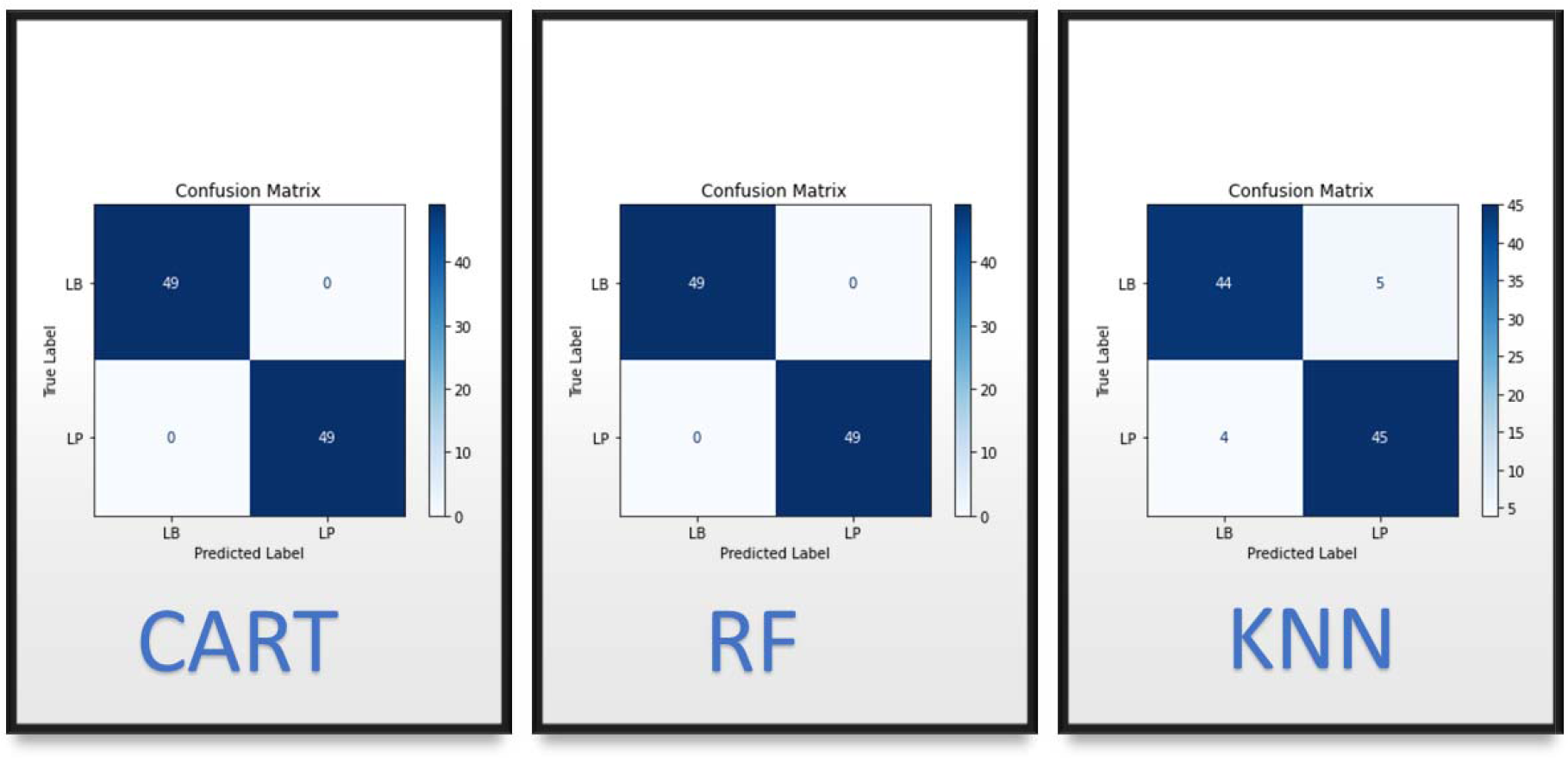
Confusion matrix scores of CART, RF, and KNN, respectively.

### 3.2. Evaluation of the Training dataset

We determined 90% accuracy in training data to be a prediction threshold. For the prediction of VOUS variants, we utilized ML methods with accuracy scores greater than 90% on training data, which were deemed successful. KNN (91%), RF (100%) and CRAT (100%) were the three ML methods that passed the threshold. The confusion matrix of these three ML techniques was depicted in Figure 3. After evaluating the training dataset, we determined that Revel scores are the most important feature, while FATHMM is the least effective. Figure 4 depicts the importance scores for characteristics.

**Figure 3.**
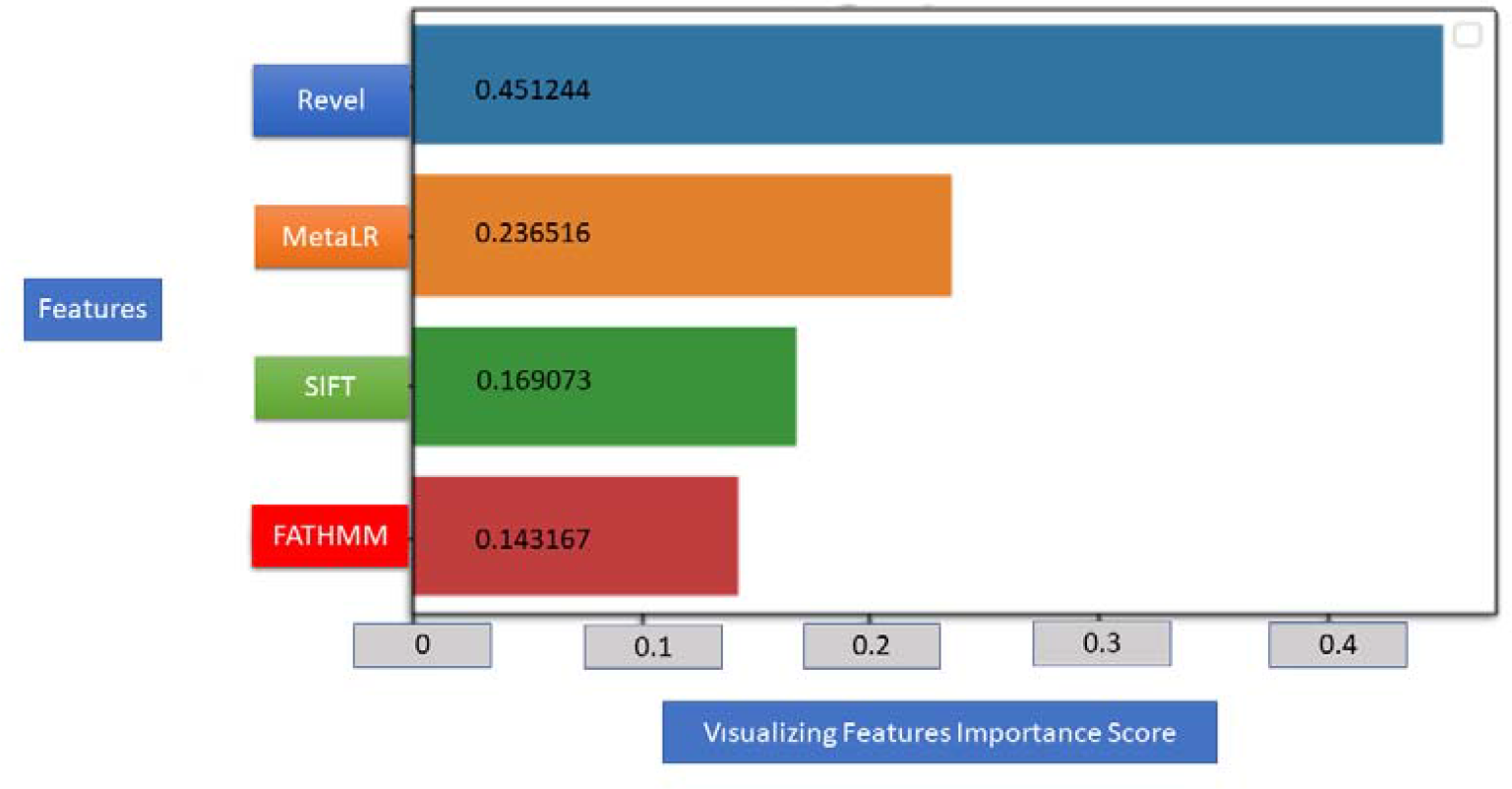
Visualizing Feature importance Scores

### 3.3. Prediction Outcomes and Evaluation of Machine Learning Algorithms on VOUS variants

We weighted the values of each successful ML method as if they predicted LP for 1 and LB for - 1. If the overall score was greater than 0, we classified the variant as LP; otherwise, it was classified as LB. For this evaluation, we found 85 LP variants and 83 LB variants. As a result, we discovered 134 LP variants and 132 LB variants in the overall dataset. Supplementary Table 1 displays the results of our scoring system.

### 3.4. Functional Evaluation

#### 3.4.1. Gene-level (Exonic) Ascertainment

First, we analyzed predicted VOUS variants. While 32 of 85 LP predicted variants were located in exon 10, 34 of 83 LB predicted variant was found in exon 2. Our prediction method concluded that 32 of 52 variants in exon 10 were LP, and 34 of 58 exon 2 variants were LB. Afterwards, we merged our prediction results with train datasets (LB and LP) and evaluated them for exonic positions [Table 1]. Thus, we obtained 134 LP and 132 LB variants. We detected LP variants much higher than LBs in exons 7,9,10, and for other exons, vice versa. While 57 of 134 LP variants were located in exon 10, 57 of 132 LB variants were positioned in exon 2. We compared LB and LP distributions and detected a statistically significant difference in exon 2 and exon 10. According to this, while exon 10 variants were about 2.6 times more likely to BE LP than LB (χ2 :12.858, p < 0.001, OR: 2.629 [1.539-4.493]), exon 2 variants with the same ratio of exon 10, but conversely more likely to be LB than LP (χ2 :12.693, p < 0.001, OR: 2.595 [1.532-4.132]).

**Table 1.** Distribution of all variants by exons and variant prediction outcomes

| Exons | Variant prediction outcomes |  |  | Odds ratios |  |  |
| --- | --- | --- | --- | --- | --- | --- |
|  | LB (n=132) | LP (n=134) | p values | Beta Values | Lower | Upper |
| 1 | 4 | 11 | 0.118<sup>a</sup> | 2.862 | 0.887 | 9.228 |
| 2 | 57 | 30 | <0.001<sup>b</sup> | 0.380 | 0.223 | 0.646 |
| 3 | 20 | 14 | 0.334<sup>a</sup> | 0.653 | 0.315 | 1.356 |
| 4 | 3 | 2 | 0.683<sup>c</sup> | 0.652 | 0.107 | 3.963 |
| 5 | 14 | 10 | 0.463<sup>a</sup> | 0.680 | 0.291 | 1.590 |
| 6 | - | - | * | * | * | * |
| 7 | - | 4 | 0.122<sup>c</sup> | * | * | * |
| 8 | 1 | 4 | 0.370<sup>c</sup> | 4.031 | 0.445 | 36.549 |
| 9 | 4 | 2 | 0.445<sup>c</sup> | 0.485 | 0.087 | 2.693 |
| 10 | 29 | 57 | <0.001<sup>b</sup> | 2.629 | 1.539 | 4.493 |

a Chi-square(Yates correction), b. Pearson chi-square test, c. Fisher Exact test,\*not calculated.

#### 3.4.2. Protein-Level (Domain-based) Evaluation

In domains, we identified 40 of 85 (47%) predicted LP variants and 24 of 83 (28.92%) predicted LB variants. When evaluating the predicted variants (n=168), domain-located variants were 2,766 times more likely to be LP than LB (χ2:10.566, p:0.002, OR: 2.766 [1.462-5.233]) After that, we merged the training dataset into the Predicted VOUS variants. We evaluated all variants collectively and discovered that LP variants were approximately 2.5 times more prevalent in domains than LB variants (χ2:13.574, p < 0.001, OR: 2.509 [1.532-4.132]). After detecting this statistically significant difference, we evaluated all variants for their specific domains [Table 2]. I In contrast, B30.2 domain variants were 2.5 times more likely to be LP than LB (χ2:12.693, p < 0.001, OR: 2.595 [1.532-4.132]). However, variants that were not detected in any domains were 2.6 times more likely to be LB than LP (χ2:14.508, p < 0.001, OR: 0.386 [0.235-0.633]).

**Table 2.**
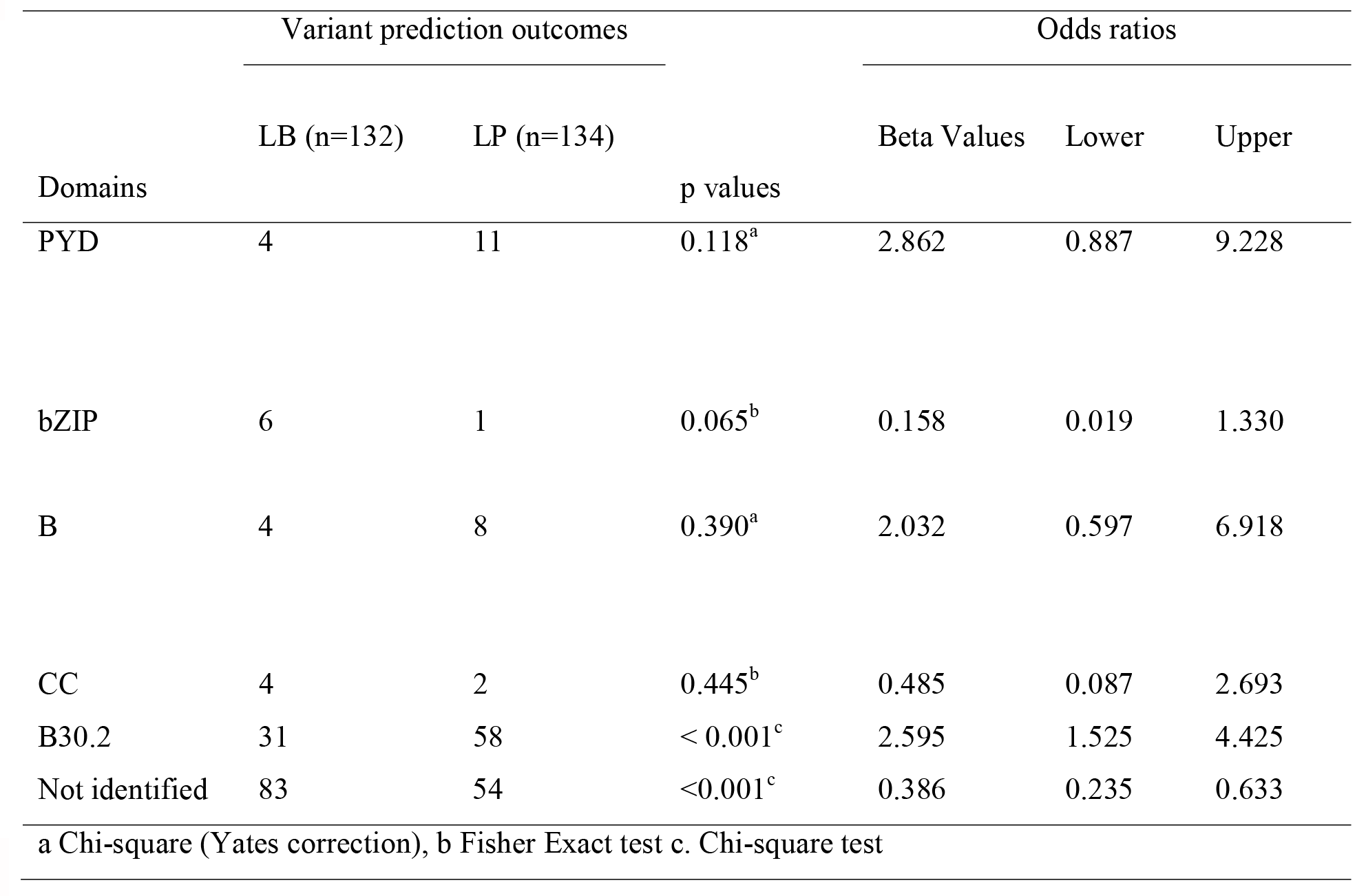
Distribution of all variants by domains and variant prediction outcomes

### 3.5. Clinical Evaluation

FMF has a complex inheritance pattern, and genetically heterogenous, therefore it was not easy to assess clinical variants^1,26^. Hence, we conducted a clinical evaluation for only consensus 14 variants^25^. The most prevalent and pathogenic seven variants (M694I, M680I, V726A, R761H, A744S, E167D, T267I) classified all three ML methods with 100 % accuracy. Clinical evaluation of 5 VOUS variants (K695R, E148Q, P369S, F479L ve I591T) classification results, we searched the literature and clinical databases. While E148Q evaluated LB for 2 ML methods, and one for LP; P369S evaluated LP for 2 ML methods, and one for LB. All ML methods evaluated K695R and F749L variants as LP, and I591T was classified as an LB. The clinical impact of E148Q variants were contradictory^27^, however, most experts’ opinion were E148Q mutations were not pathogenic^3,7,28^. The last studies supported this idea. For this, few E148Q variants with clinical manifestations generally occur compound heterozygous state^29^. Variant prediction tools (varsome, franklin) compatible with this opinion accepted this variant as benign. Although the homozygous P369S variant may show variable expression, it usually presents clinical findings which expected abdominal pain, chest pain, and fever^7,28,30,31^. K695R mutations were most likely associated with arthritis^28,30,32–34^. Studies indicated that K695R mutations were more prevalent in FMF with inflammatory bowel diseases than in isolated FMF patients^35^. FMF patients with F479L mutations, which was rarer than others, generally present with clinical manifestations, one patient with F479L mutations in homozygous state displays pleuritic chest pain, and after initiation of colchicine treatment no symptoms occur^30^. I591T variants, just like the F479L variants^30^, were observed as very rare and few reports are available in the literature^31^. Clinical outcomes of I591T variants were most likely based on previous studies, and in these studies, these variants were most likely evaluated as polymorphism or responsible for the milder presentation of the diseases^36^. Therefore, our algorithms’ outcomes were compatible with the existing literature.

## 4. DISCUSSION

In our study, we evaluated all variants in three states: First, we implemented a machine learning algorithms on the training set (LP and LB variants). To predict unknown significance variants, we then selected three of the seven machine learning methods with the highest classification accuracy (CART: 100%, RT: 100%, and KNN: 91%) (CART: 100%, RT: 100%, and KNN: 91%). After that, we added VOUS variant prediction results to previously identified LB/LP variants and evaluated all variants from a functional and clinical standpoint. We carried out functional level evaluation in two steps, at the gene and protein levels. We revealed that while pathogenic variants were more prevalent in exon 10, the opposite was true for the other genes, with the exception of exons 7 and 9, in our gene-level analysis.We found that LP variants were three times more likely to be distributed in functional domains and hotspot regions of the MEFV gene than LB variants that are compatible with ACMG recommendations, in our protein-level ascertainment. In the finat step, we implemented a clinical evaluation. This evaluation indicates that our algorithm discriminates consensus 14 pathogenic and benign variants with 100% accuracy and might provide a convenient approach for the interpretation of VOUS variants.

This study contains a number of enhanced methods and novel applications. First, our novel approach is based on three-step: (1) Implementing machine learning methods (2) Functional level evaluation ascertaintment and (3) Clinical evaluation. Second, our methodology is not founded on a single protein prediction tool or meta-predictor. Third, we did not evaluated variants using a single score, we evaluated variants with a novel ensembl method. Fourth, we implemented our ensembl method while using hard-margin which is more robust than soft margin. Furthermore, we determined the threshold for training data accuracy to be 90%. ML methods ranked higher than this criterion were accepted as successful and used for predicting VOUS variants. While the sample size based on the reported number of variants is the most important limitation of our study, we made an attempt to mitigate this by increasing the number of prediction tools and meta-predictors. While we selected prediction programs, we restricted the number of tools to the optimum level and excluded any tools with missing values. The second limitation is that our predictions are based on ML methods, and the outcomes of our study need to be confirmed in functional studies. The third limitation of our study is our clinical evaluation of variants restricted to consensus 14 variants.

First, we extracted MEFV variants from infevers databases, the most frequently employed autoinflammatory databases^37^. After extracting data from infever’s database, the first necessary step was to determine the optimal number of tools for variant prediction. There are numerous meta-predictors and amino acid alteration pathogenicity prediction scores^12^currently available. Nevertheless, we selected internationally recognized databases and implemented in silico prediction programs to assess variants. For this reason, we chose the prediction programs used in the prediction algorithm of both varsome^40^ and franklin genoox^41^. In addition, we sought the condition that there should be no missing data in selecting the score to be used. Finally, we evaluated which scores made the most effective classification when evaluating rare variants in the studies. According to previous studies, the Revel score made the most appropriate prediction for variants with an Allele frequency (AF) of 3% and below, and the MetaLR score for variants with 3-5% of AF^23^. Therefore, we presumed that these two scores would have the highest predictive power for identifying these MEFV gene variants. In light of this, we intended to incorporate these two scores into our prediction algorithm. Then, we determined if the two scores had a multicollinearity issue. There was no evidence of multicollinearity between the two metapredictors, so we included them in the algorithm. Similarly, Revel and MetaLR scores algorithms included SIFT and FATHMM scores among their prediction scores. However, no multicollinearity problems were detected between the meta-predictors included and the amino acid prediction scores. Similar approach was used previous studies^38^. Afterwards, after the outlier detection, no extreme value could be detected that would affect the result. Therefore, all variables were included in the analysis.

It is highly recommended that, when developing novel algorithms, one should not only concentrate on novelty but also identify a good feature dataset^39^. Therefore, we examined our training dataset and determined the threshold for 90% accuracy required for machine learning algorithms to succeed. This way, we only evaluated ML algorithms with higher accuracy scores than 90 percent of those selected and included in the evaluation of our prediction, excluding all others. Three machine learning (ML) algorithms, KNN (91%), RF (100%), and CRAT (100%), passed this threshold. Implementing three ML algorithms has more benefits than achieving two scores of 100 percent accuracy. Thus, indeterminacies resulting from the evaluation of two algorithms are prevented. In the next step, we used the hard-margin method, which is more robust than the soft-margin method, to obtain more accurate overall prediction score results. A few years ago, Accetturo et al.^9^ implemented 5 ML methods on 216 *MEFV* missense variants available in infevers databases to discriminate pathogenic and benign variants, and they found LDA had the most successful ML algorithm with 75 % of accuracy. They only used Revel score for predicting MEFV variants pathogenicity. Nevertheless, our study revealed that Revel scores, the most contributed amino acid prediction score, had a pathogenicity prediction accuracy of 42% out of all importance scores [Figure 3]. Our triumph of high training data accuracy score stem from optimum number of tools. Similarly, another success of our study on Accetturo et al.^9^, while their prediction was only based on most succesfull ML algorithms, our prediction was based on ensemble method.

Our initial predictions are based on ML techniques. Therefore, it must be validated by functional and clinical studies. The most effective method for validating novel variants is to conduct functional studies (BS3 and PS3, per ACMG) ^14,42^. Nevertheless, designing and conducting functional studies for 168 VOUS variants in the MEFV gene is a time- and cost-intensive endeavor ^39,43^. As a result, we performed our functional ascertainment^11,44^, based on the fact that the variant was located in a hotspot region or a well-known domain (PM1 per ACMG)^44^. Our model predicts a statistically significant difference in pathogenicity between domains and other regions, despite the fact that it lacks prior information on variant location on domains or motifs.

According to our model outcomes, LP variants are 2.5 times more likely to be located in domains than LB variants, which contradicts the findings of Accetturo et al.^9^, who discovered random distribution of LB and LP mutations. Recent studies, however, indicate distributions of pathogenic variants compatible with particular domains, such as the majority of exon 10 pathogenic variants being in SPRY domains^7,45^. In our study, SPRY domain mutations were 2.5 times more likely to be pathogenic than benign. It is known that domains are a specific physical region or amino acid sequence in a protein that is associated with a particular function or corresponding segment of DNA, and that disease-causing mutations most likely arise from mutations in these regions; thus, it is possible that domains are more likely than other protein locations to contain pathogenic mutations. In addition, we found more pathogenic mutations than benign ones in exon 10. This occurrence is most likely related to the exone responsible for the formation of the C-terminal B30.2 domain (SPRY). Variants of the SPRY gene were 2,6 times more likely to be disease-causing than the neutral form. In the final step, consensus 14 variants for clinical evaluation were evaluated. We compared our redefined variants to existing scientific literature (PubMed, MEDLINE, Scopus, Web of Science) and clinical databases (Clinvar, Intervar). Our prediction algorithms correctly classified 9 of 14 LP/P variants with a 100 percent accuracy rate. The results of the other five VOUS variants were consistent with clinical databases and previous studies^28,30,32,33,35^. However, more information on these five variants in the literature is required for more accurate predictions; novel clinical data for these variants must be collected.

Consequently, among the machine learning methods used to classify MEFV variants, our classification method yielded the most accurate results on training datasets. Our current machine learning algorithm correctly classified all (100%) training dataset variants. The fact that disease-causing MEFV variants are frequently located in domains improves the accuracy of our algorithm’s predictions. Moreover, our clinical evaluation outcomes are consistent with those of previous studies. Considering that 60% of the clinical effects of MEFV gene variants are unresolved, the evaluation of our methods in conjunction with the clinical manifestations of patients significantly simplifies the interpretation of unknown variants. To determine the success of our method in predicting the expression of other genes, however, additional research on a variety of genes is required.

## Data Availability

All data produced in the present study are available upon reasonable request to the authors

https://infevers.umai-montpellier.fr/web/search.php?n=1

## Acknowledgements

Authors thanks to Dr.Atakan Şahin for his helping in data collection step.

## Web resources

ClinVar, https://www.ncbi.nlm.nih.gov/clinvar/

DbNFSP, http://database.liulab.science/dbNSFP

Infevers, https://infevers.umai-montpellier.fr/web/

Franklin, https://franklin.genoox.com/clinical-db/home

FATHMM, http://fathmm.biocompute.org.uk/

Polyphen-2, http://genetics.bwh.harvard.edu/pph2/

REVEL, https://sites.google.com/site/revelgenomics/?pli=1

SIFT, https://sift.bii.a-star.edu.sg/

Swiss-Prot, https://www.uniprot.org/

Variant Effect Predictor, https://www.ensembl.org/Tools/VEP

Varsome, https://varsome.com/

