## Supplementary Table 1 for "THREE STEPS NOVEL HARD MARGIN ENSEMBLE MACHINE LEARNING METHOD CLASSIFIES UNCERTAIN *MEFV* GENE VARIANTS"

| HGVS sequence name | HGVS protein name | Usual name | Infevers Classification | CART | RF | KNN | Our Classification Results |
| --- | --- | --- | --- | --- | --- | --- | --- |
| c.2333A>T | p.(Gln778Leu) | Q778L | VOUS | LP | LB | LB | LB |
| c.289C>A | p.(Gln97Lys) | Q97K | VOUS | LB | LB | LB | LB |
| c.290A>G | p.(Gln97Arg) | Q97R | VOUS | LP | LP | LP | LP |
| c.311C>G | p.(Ser104Cys) | S104C | NC | LP | LP | LP | LP |
| c.314C>A | p.(Ala105Glu) | A105E | VOUS | LB | LB | LB | LB |
| c.322A>C | p.(Ser108Arg) | S108R | VOUS | LB | LB | LP | LB |
| c.322A>G | p.(Ser108Gly) | S108G | VOUS | LB | LB | LB | LB |
| c.329T>C | p.(Leu110Pro) | L110P | VOUS | LB | LB | LB | LB |
| c.331G>A | p.(Gly111Arg) | G111R | VOUS | LP | LB | LB | LB |
| c.332G>A | p.(Gly111Glu) | G111E | VOUS | LB | LB | LB | LB |
| c.344C>G | p.(Pro115Arg) | P115R | VOUS | LB | LB | LB | LB |
| c.369C>A | p.(His123Gln) | H123Q | VOUS | LB | LB | LB | LB |
| c.406G>A | p.(Gly136Arg) | G136R | VOUS | LP | LP | LP | LP |
| c.406G>T | p.(Gly136Trp) | G136W | VOUS | LP | LP | LP | LP |
| c.407G>A | p.(Gly136Glu) | G136E | VOUS | LP | LB | LB | LB |
| c.433A>G | p.(Ser145Gly) | S145G | VOUS | LB | LB | LB | LB |
| c.442G>C | p.(Glu148Gln) | E148Q | VOUS | LB | LB | LB | LB |
| c.443A>T | p.(Glu148Val) | E148V | VOUS | LP | LP | LP | LP |
| c.444G>C | p.(Glu148Asp) | E148D | VOUS | LP | LP | LB | LP |
| c.453G>C | p.(Arg151Ser) | R151S | VOUS | LB | LB | LB | LB |
| c.454G>C | p.(Gly152Arg) | G152R | VOUS | LP | LP | LP | LP |
| c.460T>C | p.(Ser154Pro) | S154P | VOUS | LB | LB | LB | LB |
| c.464G>C | p.(Arg155Thr) | R155T | VOUS | LB | LB | LB | LB |
| c.488A>C | p.(Glu163Ala) | E163A | VOUS | LB | LB | LB | LB |
| c.489G>T | p.(Glu163Asp) | E163D | Unsolved | LP | LB | LB | LB |
| c.493G>A | p.(Ala165Thr) | A165T | VOUS | LB | LB | LB | LB |
| c.497C>T | p.(Ser166Leu) | S166L | VOUS | LP | LB | LB | LB |
| c.515A>C | p.(Gln172Pro) | Q172P | VOUS | LP | LP | LP | LP |
| c.524C>A | p.(Pro175His) | P175H | VOUS | LB | LB | LB | LB |
| c.536G>T | p.(Ser179Ile) | S179I | VOUS | LB | LB | LP | LB |
| c.539C>G | p.(Pro180Arg) | P180R | VOUS | LP | LP | LB | LP |
| c.547C>A | p.(Pro183Thr) | P183T | VOUS | LP | LB | LB | LB |
| c.560G>A | p.(Ser187Asn) | S187N | NC | LB | LB | LB | LB |
| c.578C>T | p.(Ala193Val) | A193V | Unsolved | LB | LB | LB | LB |
| c.586G>A | p.(Gly196Arg) | G196R | VOUS | LB | LB | LB | LB |
| c.586G>T | p.(Gly196Trp) | G196W | VOUS | LB | LB | LP | LB |
| c.589G>C | p.(Gly197Arg) | G197R | VOUS | LP | LB | LP | LP |
| c.608T>C | p.(Leu203Pro) | L203P | VOUS | LB | LB | LP | LB |
| c.611G>A | p.(Arg204His) | R204H | NC | LP | LP | LP | LP |
| c.617A>G | p.(Asn206Ser) | N206S | VOUS | LP | LP | LP | LP |
| c.618C>G | p.(Asn206Lys) | N206K | VOUS | LP | LP | LB | LP |
| c.622A>T | p.(Ser208Cys) | S208C | VOUS | LP | LP | LP | LP |
| c.653G>C | p.(Gly218Ala) | G218A | VOUS | LB | LB | LB | LB |
| c.664G>A | p.(Gly222Arg) | G222R | VOUS | LB | LP | LP | LP |
| c.674A>G | p.(Glu225Gly) | E225G | VOUS | LP | LP | LP | LP |
| c.675G>C | p.(Glu225Asp) | E225D | Unsolved | LB | LB | LP | LB |
| c.688G>A | p.(Glu230Lys) | E230K | VOUS | LB | LB | LB | LB |
| c.704C>T | p.(Ser235Leu) | S235L | NC | LP | LP | LP | LP |
| c.707G>T | p.(Gly236Val) | G236V | Unsolved | LP | LP | LP | LP |
| c.714G>T | p.(Met238Ile) | M238I | VOUS | LB | LB | LB | LB |
| c.730G>A | p.(Glu244Lys) | E244K | NC | LP | LP | LP | LP |
| c.745A>G | p.(Thr249Ala) | T249A | VOUS | LB | LB | LB | LB |
| c.751G>A | p.(Glu251Lys) | E251K | VOUS | LP | LP | LP | LP |
| c.770C>T | p.(Pro257Leu) | P257L | VOUS | LP | LP | LP | LP |
| c.848C>G | p.(Pro283Arg) | P283R | VOUS | LP | LB | LB | LB |
| c.848C>T | p.(Pro283Leu) | P283L | VOUS | LP | LP | LP | LP |
| c.863C>A | p.(Ser288Tyr) | S288Y | VOUS | LB | LB | LB | LB |
| c.866C>A | p.(Ala289Glu) | A289E | VOUS | LP | LP | LP | LP |
| c.866C>T | p.(Ala289Val) | A289V | VOUS | LP | LP | LP | LP |
| c.1012G>C | p.(Val338Leu) | V338L | VOUS | LB | LB | LB | LB |
| c.1016C>T | p.(Ser339Phe) | S339F | Unsolved | LP | LP | LP | LP |
| c.1043G>A | p.(Arg348His) | R348H | Unsolved | LB | LB | LB | LB |
| c.1048C>T | p.(Pro350Ser) | P350S | VOUS | LP | LP | LP | LP |
| c.1049C>G | p.(Pro350Arg) | P350R | VOUS | LB | LB | LB | LB |
| c.1057C>G | p.(Pro353Ala) | P353A | NC | LP | LB | LB | LB |
| c.1082G>C | p.(Arg361Thr) | R361T | VOUS | LP | LP | LB | LP |
| c.1088G>A | p.(Ser363Asn) | S363N | VOUS | LP | LB | LB | LB |
| c.1091C>T | p.(Pro364Leu) | P364L | VOUS | LB | LB | LB | LB |
| c.1099C>G | p.(Leu367Val) | L367V | NC | LB | LB | LP | LB |
| c.1105C>T | p.(Pro369Ser) | P369S | VOUS | LP | LB | LP | LP |
| c.749G>C | p.(Gly250Ala) | G250A | VOUS | LB | LB | LB | LB |
| c.926C>T | p.(Thr309Met) | T309M | VOUS | LB | LB | LB | LB |
| c.940C>T | p.(Arg314Cys) | R314C | VOUS | LB | LB | LB | LB |
| c.941G>A | p.(Arg314His) | R314H | Unsolved | LB | LB | LB | LB |
| c.955G>A | p.(Glu319Lys) | E319K | Unsolved | LB | LP | LB | LB |
| c.983T>C | p.(Val328Ala) | V328A | Unsolved | LB | LB | LB | LB |
| c.1324T>A | p.(Ser442Thr) | S442T | VOUS | LB | LB | LB | LB |
| c.1368A>C | p.(Glu456Asp) | E456D | Unsolved | LP | LP | LP | LP |
| c.1420G>A | p.(Glu474Lys) | E474K | VOUS | LP | LP | LP | LP |
| c.1432C>T | p.(His478Tyr) | H478Y | VOUS | LB | LB | LB | LB |
| c.1450C>A | p.(Leu484Met) | L484M | NC | LP | LP | LB | LP |
| c.1459G>A | p.(Val487Met) | V487M | VOUS | LB | LB | LB | LB |
| c.1459G>C | p.(Val487Leu) | V487L | Unsolved | LB | LB | LB | LB |
| c.1484G>A | p.(Arg495Lys) | R495K | VOUS | LB | LB | LB | LB |
| c.1501C>G | p.(Arg501Gly) | R501G | VOUS | LP | LP | LB | LP |
| c.1514A>G | p.(Asp505Gly) | D505G | VOUS | LP | LP | LB | LP |
| c.1523T>A | p.(Leu508Gln) | L508Q | VOUS | LP | LP | LP | LP |
| c.1528G>A | p.(Asp510Asn) | D510N | VOUS | LB | LB | LP | LB |
| c.1532C>A | p.(Ala511Glu) | A511E | VOUS | LB | LB | LB | LB |
| c.1532C>T | p.(Ala511Val) | A511V | VOUS | LB | LB | LB | LB |
| c.1538T>C | p.(Ile513Thr) | I513T | Unsolved | LP | LP | LP | LP |
| c.1541G>A | p.(Gly514Glu) | G514E | VOUS | LB | LB | LB | LB |
| c.1559A>T | p.(Glu520Val) | E520V | NC | LB | LP | LB | LB |
| c.1617G>T | p.(Lys539Asn) | K539N | VOUS | LP | LP | LP | LP |
| c.1648C>G | p.(Pro550Ala) | P550A | VOUS | LP | LP | LP | LP |
| c.1656G>C | p.(Glu552Asp) | E552D | VOUS | LP | LP | LB | LP |
| c.1675C>T | p.(Leu559Phe) | L559F | Unsolved | LP | LP | LP | LP |
| c.1772T>C | p.(Ile591Thr) | I591T | VOUS | LB | LB | LB | LB |
| c.1784C>T | p.(Ala595Val) | A595V | VOUS | LB | LP | LP | LP |
| c.343C>A | p.(Pro115Thr) | P115T | LB | LB | LB | LB | LB |
| c.359C>T | p.(Thr120Ile) | T120I | LB | LB | LB | LB | LB |
| c.422G>T | p.(Ser141Ile) | S141I | LB | LB | LB | LP | LB |
| c.428G>C | p.(Arg143Pro) | R143P | LB | LB | LB | LP | LB |
| c.476G>A | p.(Ser159Asn) | S159N | LB | LB | LB | LB | LB |
| c.505C>G | p.(Leu169Val) | L169V | LB | LB | LB | LB | LB |
| c.530C>T | p.(Thr177Ile) | T177I | LB | LB | LB | LB | LB |
| c.536G>A | p.(Ser179Asn) | S179N | LB | LB | LB | LB | LB |
| c.581T>C | p.(Leu194Pro) | L194P | LB | LB | LB | LB | LB |
| c.605G>A | p.(Arg202Gln) | R202Q | B | LB | LB | LB | LB |
| c.688G>C | p.(Glu230Gln) | E230Q | LB | LB | LB | LP | LB |
| c.694T>C | p.(Tyr232His) | Y232H | LB | LB | LB | LB | LB |
| c.722G>A | p.(Arg241Lys) | R241K | LB | LB | LB | LB | LB |
| c.739A>G | p.(Ile247Val) | I247V | LB | LB | LB | LB | LB |
| c.775A>G | p.(Ile259Val) | I259V | LB | LB | LB | LB | LB |
| c.833G>C | p.(Arg278Pro) | R278P | LB | LB | LB | LB | LB |
| c.896A>G | p.(Glu299Gly) | E299G | LB | LB | LB | LP | LB |
| c.910G>A | p.(Gly304Arg) | G304R | LB | LB | LB | LB | LB |
| c.949G>A | p.(Ala317Thr) | A317T | LB | LB | LB | LB | LB |
| c.986G>A | p.(Arg329His) | R329H | LB | LB | LB | LB | LB |
| c.1006G>A | p.(Glu336Lys) | E336K | LB | LB | LB | LB | LB |
| c.1370C>T | p.(Ala457Val) | A457V | LB | LB | LB | LB | LB |
| c.1382G>A | p.(Arg461Gln) | R461Q | LB | LB | LB | LB | LB |
| c.1405G>T | p.(Val469Leu) | V469L | LB | LB | LB | LB | LB |
| c.1502G>A | p.(Arg501His) | R501H | LB | LB | LB | LB | LB |
| c.1516A>G | p.(Ile506Val) | I506V | LB | LB | LB | LB | LB |
| c.2338C>A | p.(Pro780Thr) | P780T | LB | LB | LB | LB | LB |
| c.115A>G | p.(Arg39Gly) | R39G | LP | LP | LP | LP | LP |
| c.250G>A | p.(Glu84Lys) | E84K | LP | LP | LP | LP | LP |
| c.265G>A | p.(Ala89Thr) | A89T | LP | LP | LP | LP | LP |
| c.501G>C | p.(Glu167Asp) | E167D | LP | LP | LP | LP | LP |
| c.623G>C | p.(Ser208Thr) | S208T | LP | LP | LP | LP | LP |
| c.724A>G | p.(Ser242Gly) | S242G | LP | LP | LP | LP | LP |
| c.726C>A | p.(Ser242Arg) | S242R | LP | LP | LP | LP | LP |
| c.726C>G | p.(Ser242Arg) | S242R | LP | LP | LP | LP | LP |
| c.800C>T | p.(Thr267Ile) | T267I | LP | LP | LP | LP | LP |
| c.938C>A | p.(Pro313His) | P313H | LP | LP | LP | LP | LP |
| c.1060C>T | p.(Arg354Trp) | R354W | LP | LP | LP | LP | LP |
| c.1437C>G | p.(Phe479Leu) | F479L | LP | LP | LP | LP | LP |
| c.1501C>T | p.(Arg501Cys) | R501C | LP | LP | LP | LP | LP |
| c.1508C>G | p.(Ser503Cys) | S503C | LP | LP | LP | LP | LP |
| c.1729A>G | p.(Thr577Ala) | T577A | LP | LP | LP | LP | LP |
| c.1729A>T | p.(Thr577Ser) | T577S | LP | LP | LP | LP | LP |
| c.1730C>A | p.(Thr577Asn) | T577N | LP | LP | LP | LB | LP |
| c.1730C>G | p.(Thr577Ser) | T577S | LP | LP | LP | LP | LP |
| c.101A>C | p.(Gln34Pro) | Q34P | NC | LB | LB | LB | LB |
| c.104A>G | p.(Lys35Arg) | K35R | NC | LB | LB | LP | LB |
| c.124C>T | p.(Arg42Trp) | R42W | Unsolved | LP | LB | LP | LP |
| c.149C>T | p.(Pro50Leu) | P50L | NC | LP | LP | LP | LP |
| c.184G>T | p.(Gly62Trp) | G62W | NC | LP | LP | LP | LP |
| c.200T>A | p.(Val67Glu) | V67E | VOUS | LP | LP | LP | LP |
| c.35C>T | p.(Thr12Ile) | T12I | Unsolved | LP | LP | LP | LP |
| c.56A>G | p.(Tyr19Cys) | Y19C | Unsolved | LP | LP | LP | LP |
| c.82C>G | p.(Leu28Val) | L28V | VOUS | LP | LP | LP | LP |
| c.224G>A | p.(Arg75Gln) | R75Q | LB | LB | LB | LB | LB |
| c.260A>G | p.(His87Arg) | H87R | B | LB | LB | LB | LB |
| c.74A>G | p.(Lys25Arg) | K25R | LP | LP | LP | LP | LP |
| c.796A>G | p.(Lys266Glu) | K266E | LB | LB | LB | LB | LB |
| c.803C>T | p.(Ala268Val) | A268V | LB | LB | LB | LB | LB |
| c.808A>G | p.(Asn270Asp) | N270D | LB | LB | LB | LB | LB |
| c.818C>T | p.(Ser273Leu) | S273L | LB | LB | LB | LB | LB |
| c.829C>A | p.(Pro277Thr) | P277T | LB | LB | LB | LB | LB |
| c.1118C>T | p.(Pro373Leu) | P373L | VOUS | LP | LB | LP | LP |
| c.1147C>A | p.(Gln383Lys) | Q383K | VOUS | LP | LP | LB | LP |
| c.1211A>G | p.(His404Arg) | H404R | NC | LP | LP | LP | LP |
| c.1223G>A | p.(Arg408Gln) | R408Q | VOUS | LB | LB | LB | LB |
| c.1229G>A | p.(Arg410His) | R410H | NC | LP | LB | LB | LB |
| c.1127A>G | p.(Lys376Arg) | K376R | LB | LB | LB | LB | LB |
| c.1171G>A | p.(Asp391Asn) | D391N | LB | LB | LB | LB | LB |
| c.1115T>C | p.(Leu372Pro) | L372P | LP | LP | LP | LB | LP |
| c.1151T>C | p.(Leu384Pro) | L384P | LP | LP | LP | LP | LP |
| c.1166A>T | p.(Asp389Val) | D389V | LP | LP | LP | LP | LP |
| c.1186C>T | p.(Leu396Phe) | L396F | LP | LP | LP | LP | LP |
| c.1207G>A | p.(Glu403Lys) | E403K | LP | LP | LP | LP | LP |
| c.1267A>G | p.(Ile423Val) | I423V | VOUS | LP | LP | LP | LP |
| c.1268T>C | p.(Ile423Thr) | I423T | NC | LP | LP | LP | LP |
| c.1277A>G | p.(Gln426Arg) | Q426R | VOUS | LB | LB | LP | LB |
| c.1318C>G | p.(Gln440Glu) | Q440E | LB | LB | LB | LB | LB |
| c.1795A>G | p.(Asn599Asp) | N599D | VOUS | LB | LB | LB | LB |
| c.1876T>A | p.(Trp626Arg) | W626R | NC | LB | LB | LB | LB |
| c.1894G>A | p.(Gly632Ser) | G632S | Unsolved | LP | LB | LB | LB |
| c.1898C>T | p.(Pro633Leu) | P633L | NC | LP | LP | LP | LP |
| c.1907T>A | p.(Phe636Tyr) | F636Y | NC | LP | LP | LP | LP |
| c.1910A>G | p.(Asp637Gly) | D637G | Unsolved | LP | LP | LB | LP |
| c.1920C>G | p.(Ile640Met) | I640M | VOUS | LB | LB | LB | LB |
| c.1921A>T | p.(Ile641Phe) | I641F | VOUS | LB | LB | LB | LB |
| c.1925T>C | p.(Val642Ala) | V642A | NC | LB | LB | LB | LB |
| c.1937C>T | p.(Pro646Leu) | P646L | VOUS | LB | LB | LB | LB |
| c.1945C>T | p.(Leu649Phe) | L649F | NC | LB | LB | LB | LB |
| c.1946T>C | p.(Leu649Pro) | L649P | VOUS | LP | LB | LP | LP |
| c.1954C>T | p.(Arg652Cys) | R652C | VOUS | LB | LP | LB | LB |
| c.1955G>A | p.(Arg652His) | R652H | VOUS | LP | LP | LP | LP |
| c.1957C>A | p.(Arg653Ser) | R653S | VOUS | LB | LB | LB | LB |
| c.1958G>A | p.(Arg653His) | R653H | VOUS | LP | LB | LB | LB |
| c.1981G>A | p.(Asp661Asn) | D661N | Unsolved | LB | LB | LB | LB |
| c.1981G>T | p.(Asp661Tyr) | D661Y | NC | LP | LP | LP | LP |
| c.2012A>T | p.(Lys671Met) | K671M | Unsolved | LP | LP | LP | LP |
| c.2033G>A | p.(Gly678Glu) | G678E | VOUS | LP | LP | LP | LP |
| c.2035A>C | p.(Asn679His) | N679H | VOUS | LP | LB | LB | LB |
| c.2042C>T | p.(Thr681Ile) | T681I | VOUS | LP | LP | LP | LP |
| c.2053G>A | p.(Glu685Lys) | E685K | VOUS | LB | LB | LB | LB |
| c.2069T>G | p.(Val690Gly) | V690G | NC | LP | LP | LP | LP |
| c.2072T>G | p.(Val691Gly) | V691G | NC | LP | LP | LP | LP |
| c.2078T>A | p.(Met693Lys) | M693K | NC | LP | LP | LP | LP |
| c.2079G>C | p.(Met693Ile) | M693I | Unsolved | LP | LP | LP | LP |
| c.2084A>T | p.(Lys695Met) | K695M | Unsolved | LB | LP | LP | LP |
| c.2094G>T | p.(Glu698Asp) | E698D | VOUS | LP | LP | LP | LP |
| c.2105C>G | p.(Ser702Cys) | S702C | VOUS | LB | LP | LP | LP |
| c.2110G>A | p.(Val704Ile) | V704I | Unsolved | LP | LP | LP | LP |
| c.2113C>T | p.(Pro705Ser) | P705S | Unsolved | LP | LP | LP | LP |
| c.2122C>T | p.(Arg708Cys) | R708C | VOUS | LP | LB | LB | LB |
| c.2126T>G | p.(Leu709Arg) | L709R | Unsolved | LP | LP | LP | LP |
| c.2149C>A | p.(Arg717Ser) | R717S | Unsolved | LP | LP | LP | LP |
| c.2150G>A | p.(Arg717His) | R717H | VOUS | LB | LP | LP | LP |
| c.2150G>T | p.(Arg717Leu) | R717L | VOUS | LP | LP | LP | LP |
| c.2160C>G | p.(Ile720Met) | I720M | VOUS | LP | LP | LP | LP |
| c.2164G>A | p.(Val722Met) | V722M | VOUS | LB | LB | LB | LB |
| c.2185A>G | p.(Ile729Val) | I729V | VOUS | LP | LP | LP | LP |
| c.2187C>G | p.(Ile729Met) | I729M | VOUS | LP | LP | LP | LP |
| c.2189C>G | p.(Ser730Cys) | S730C | Unsolved | LP | LP | LP | LP |
| c.2198A>G | p.(Asn733Ser) | N733S | Unsolved | LP | LP | LP | LP |
| c.2229C>G | p.(Phe743Leu) | F743L | Unsolved | LP | LP | LP | LP |
| c.2230G>A | p.(Ala744Thr) | A744T | Unsolved | LB | LB | LB | LB |
| c.2230G>T | p.(Ala744Ser) | A744S | VOUS | LP | LP | LP | LP |
| c.2231C>A | p.(Ala744Asp) | A744D | VOUS | LB | LB | LP | LB |
| c.2246C>G | p.(Ser749Cys) | S749C | VOUS | LP | LP | LP | LP |
| c.2261C>G | p.(Pro754Arg) | P754R | Unsolved | LP | LP | LP | LP |
| c.2263A>G | p.(Ile755Val) | I755V | VOUS | LB | LB | LB | LB |
| c.2272C>T | p.(Pro758Ser) | P758S | Unsolved | LP | LP | LP | LP |
| c.1792G>A | p.(Val598Ile) | V589I | NC | LP | LP | LB | LP |
| c.1744A>C | p.(Met582Leu) | M582L | LB | LB | LB | LP | LB |
| c.1764G>A | p.(Pro588=) | P588P | B | LB | LB | LB | LB |
| c.1773T>G | p.(Ile591Met) | I591M | LB | LB | LB | LB | LB |
| c.1776C>T | p.(Gly592=) | G592G | LB | LB | LB | LB | LB |
| c.1828A>G | p.(Asn610Asp) | N610D | LB | LB | LB | LB | LB |
| c.1883G>A | p.(Arg628Lys) | R628K | LB | LB | LB | LB | LB |
| c.1895G>C | p.(Gly632Ala) | G632A | LB | LB | LB | LB | LB |
| c.1996A>G | p.(Ile666Val) | I666V | LB | LB | LB | LB | LB |
| c.2024G>A | p.(Ser675Asn) | S675N | LB | LB | LB | LB | LB |
| c.2068G>C | p.(Val690Leu) | V690L | LB | LB | LB | LB | LB |
| c.2210G>A | p.(Arg737Lys) | R737K | LB | LB | LB | LB | LB |
| c.2314A>G | p.(Ile772Val) | I772V | LB | LB | LB | LB | LB |
| c.1949C>A | p.(Ser650Tyr) | S650Y | LP | LP | LP | LP | LP |
| c.1967A>C | p.(Glu656Ala) | E656A | LP | LP | LP | LP | LP |
| c.1975G>T | p.(Val659Phe) | V659F | LP | LP | LP | LP | LP |
| c.2002G>A | p.(Gly668Arg) | G668R | LP | LP | LP | LP | LP |
| c.2038A>C | p.(Met680Leu) | M680L | LP | LP | LP | LP | LP |
| c.2038A>G | p.(Met680Val) | M680V | LP | LP | LP | LB | LP |
| c.2040G>A | p.(Met680Ile) | M680I | P | LP | LP | LP | LP |
| c.2040G>C | p.(Met680Ile) | M680I | P | LP | LP | LP | LP |
| c.2040G>T | p.(Met680Ile) | M680I | LP | LP | LP | LP | LP |
| c.2060G>A | p.(Gly687Asp) | G687D | LP | LP | LP | LP | LP |
| c.2063A>G | p.(Tyr688Cys) | Y688C | LP | LP | LP | LP | LP |
| c.2063A>T | p.(Tyr688Phe) | Y688F | LP | LP | LP | LP | LP |
| c.2080A>T | p.(Met694Leu) | M694L | LP | LP | LP | LP | LP |
| c.2080A>G | p.(Met694Val) | M694V | P | LP | LP | LP | LP |
| c.2081T>A | p.(Met694Lys) | M694K | LP | LP | LP | LP | LP |
| c.2082G>A | p.(Met694Ile) | M694I | P | LP | LP | LP | LP |
| c.2084A>G | p.(Lys695Arg) | K695R | LP | LP | LP | LP | LP |
| c.2085G>C | p.(Lys695Asn) | K695N | LP | LP | LP | LP | LP |
| c.2177T>C | p.(Val726Ala) | V726A | P | LP | LP | LB | LP |
| c.2228T>A | p.(Phe743Tyr) | F743Y | LP | LP | LP | LP | LP |
| c.2259A>T | p.(Gln753His) | Q753H | LP | LP | LP | LP | LP |
| c.2281C>T | p.(Arg761Cys) | R761C | LP | LP | LP | LP | LP |
| c.2282G>A | p.(Arg761His) | R761H | LP | LP | LP | LP | LP |
| c.2296A>C | p.(Asn766His) | N766H | LP | LP | LP | LP | LP |
| c.2305C>G | p.(Pro769Ala) | P769A | LP | LP | LP | LP | LP |
